# Clinical evolution of COVID-19 during pregnancy at different altitudes: a population-based study

**DOI:** 10.1101/2020.09.14.20193177

**Authors:** Juan Alonso Leon-Abarca, Maria Teresa Peña-Gallardo, Jorge Soliz, Roberto Accinelli

## Abstract

**Background:** The impact of influenza and various types of coronaviruses (SARS-CoV and MERS-CoV) on pregnancy has been reported. However, the current pandemic caused by SARS-CoV-2 continues to reveal important data for understanding its behavior in pregnant women.

**Methods:** We analyzed the records of 326,586 non-pregnant women of reproductive age and 7,444 pregnant women with no other risk factor who also had a SARS-CoV-2 RT-PCR result to estimate adjusted prevalence (aP) and adjusted prevalence ratios (aPR) of COVID-19 and its requirement of hospitalization, intubation, ICU admission and case-fatality rates. Adjustment was done through Poisson regressions for age and altitude of residence and birth. Generalized binomial models were used to generate probability plots to display how each outcome varied across ages and altitudes.

**Results:** Pregnancy was independently associated with a 15% higher probability of COVID-19 (aPR: 1.15), a 116% higher probability of its following admission (aPR: 2.169) and a 127% higher probability of ICU admission (aPR: 2.275). Also, pregnancy was associated with 84.2% higher probability of developing pneumonia (aPR: 1.842) and a 163% higher probability of its following admission (aPR: 2.639). There were no significant differences in COVID-19 case-fatality rates between pregnant and non pregnant women (1.178, 95% CI: 0.68-1.67).

**Conclusion:** Pregnancy was associated with a higher probability of COVID-19, developing of pneumonia, hospitalization, and ICU admission. Our results also suggest that the risk of COVID-19 and its related outcomes, except for intubation, decrease with altitude.

## Introduction

Pneumonia in pregnant women, regardless of the etiology, is an important cause of morbidity and mortality. It is known that during pregnancy there are different anatomical and physiological changes such as diaphragm elevation due to the progressive increase in uterine dimensions, increased intra-abdominal pressure and intravascular volume, as well as decreased chest distension and immune response. As a consequence, the susceptibility to infection increases, in addition to the risk of developing complications and even respiratory failure **(1-3)**.

In 2002, SARS-CoV infected 8,098 people with a case fatality rate (CFR) of 10.5% **(4)**, while in 2012 MERS-CoV had a total of 2,519 confirmed cases and a CFR of 34.4% **(5)**. On the other hand, in 2009, cases of influenza A H1N1 were estimated at 60.8 million, and approximately 18,500 deaths were reported **(6-7)**. These diseases have been shown to be more severe in pregnant women.

We are currently facing the pandemic caused by the novel coronavirus, SARS-CoV-2, which has already exceeded 20 million cases worldwide **(8)**. Cases and deaths are increasing rapidly and we still lack specific treatment for the disease, and although rates of hospitalization and admission to the intensive care unit (ICU) are variable, health systems are already saturated.

Recent studies suggest the possibility that being pregnant may be a risk factor for SARS-CoV-2 infection and its complications **(9)**. Similarly, it was suggested that the infection rate of this virus might be lower at high-altitude (over 2,500m) **10)**. This study seeks to evaluate pregnancy and high-altitude as factors that could intervene in the risk of contracting SARS-CoV-2 infection, as well as the severity of its presentation.

## Materials and methods

### Data sources

We analyzed the records of Mexican women of reproductive age **(11)** (15-49 years) who were under clinical suspicion of COVID-19 because of a history of cough, fever, headache associated with dyspnea, arthralgia, myalgia, sore throat, runny nose, conjunctivitis or chest pain in the past 7 days. According to the protocols of the Mexican Ministry of Health, samples for SARS-CoV-2 RT-PCR are taken from every patient with respiratory distress and one out of ten mild cases (ambulatory) in the 245 respiratory monitoring units (“USMER”) distributed throughout the nation. These data are updated daily according to their “Open Data” policy, in which over a million confirmed, discarded or suspected COVID-19 patients are reported to date.

Only registries of patients without any other known risk factor recorded besides pregnancy with definite RT-PCR results (positive or negative) were included. Also, we only considered those who had their place of current residence and birth in the record. This information was used to estimate the mean altitude at birth and residence at the time of RT-PCR sampling, using an auxiliary dataset provided by the Mexican Geology Service. After the inclusion and exclusion criteria, 326,586 non-pregnant women and 7,444 pregnant women were analyzed. In this study we define COVID-19 as the individual who had a positive SARS-CoV-2 RT-PCR test and further separate the cases who had the additional diagnosis of pneumonia to evaluate the clinical evolution in detail.

### Data analysis

We applied robust Poisson regressions to estimate the adjusted prevalence (aP) and adjusted prevalence ratios (aPR) of 10 individual outcomes according to pregnancy status: COVID-19 (all cases) and pneumonia (COVID-19 pneumonia) and their requirement of hospitalization, intubation, Intensive Care Unit (ICU) admission and case-fatality ratio (CFR). Interaction between variables was considered for age+pregnancy and pregnancy+altitude to address the possible cluster effect of pregnant women located at diverse cities at diverse altitudes. Generalized binomial models using the same equations were used to generate marginal predictive probability plots to display how the probability of each outcome varied across age and altitude as a continuous spectrum. These probability plots were presented as trend lines with 95% confidence bands. The STATA 14.0 software was used for data analysis and p values <0.05 were considered statistically significant.

### Data handling and sharing statement

Processed data files used in the present study are available at request. The authors follow the STrengthening the Reporting of OBservational studies in Epidemiology (STROBE) **(12)** statement for data collection, analysis and reporting.

## Results

Summary characteristics of the study population are presented in **Table 1**. Pregnant women represented 2.23% of the study population and were on average six years younger than non-pregnant women (28.2 versus 34.9 years, p<0.0001). Pregnant women had a 11.3% higher proportion of COVID-19 compared to non-pregnant women (p<0.0001), and a following 132% higher proportion of hospital admission (p<0.0001) and 48.6% higher proportion of ICU admission (p=0.0014). Also, pregnant women had a 45% higher proportion of COVID-19 pneumonia, 20.3% higher proportion of admission, 90.7% higher ICU admission and a 37.8% lower case-fatality rate (p<0.0001) (Crude prevalences).

**Table 1.** – Summary characteristics of the study population. * Mean and standard deviation shown. N=334030.

| Categorical variables | Categories | n | % |  |
| --- | --- | --- | --- | --- |
| Women | Non-pregnant | 326,586 | 97.77 |  |
|  | Pregnant | 7,444 | 2.23 |  |
| Clinical evolution of non-pregnant patients | COVID-19 cases | 135,401 | 41.46 |  |
|  | Admission | 14,104 | 10.42 |  |
|  | Intubation | 18 | 2.09 |  |
|  | ICU admission | 51 | 5.91 |  |
|  | Case-fatality rate | 2,877 | 2.12 |  |
|  | Subset: COVID-19 pneumonia | 11,401 | 3.49 |  |
|  | Admission | 7,972 | 69.92 |  |
|  | Intubation | 759 | 9.53 |  |
|  | ICU admission | 756 | 9.49 |  |
|  | Case-fatality rate | 2,145 | 18.81 |  |
| Clinical evolution of pregnant patients | COVID-19 cases | 3,434 | 46.13 |  |
|  | Admission | 833 | 24.26 |  |
|  | Intubation | 42 | 5.05 |  |
|  | ICU admission | 80 | 9.63 |  |
|  | Case-fatality rate | 58 | 1.69 |  |
|  | Subset: COVID-19 pneumonia | 377 | 5.06 |  |
|  | Admission | 317 | 84.08 |  |
|  | Intubation | 36 | 11.43 |  |
|  | ICU admission | 57 | 18.1 |  |
|  | Case-fatality rate | 44 | 11.67 |  |
| Age groups percentiles (pregnant women) |  | 20 years | 10 |  |
|  |  | 24 years | 25 |  |
|  |  | 28 years | 50 |  |
|  |  | 33 years | 75 |  |
|  |  | 36 years | 90 |  |
| Continuous variables |  | n | Median (IQR) | p |
| Age * | Non-pregnant | 326,586 | 34.9 (8.5) | 0 |
|  | Pregnant | 7,444 | 28.2 (6.2) |  |
| Days from symptom onset to healthcare consult | Non-pregnant | 326,586 | 3 (3) | 0 |
|  | Pregnant | 7,444 | 3 (4) |  |
| Days from admission to death | Non-pregnant | 3683 | 5 (8) | 0.3881 |
|  | Pregnant | 81 | 7 (10) |  |

Most of pregnant women who developed COVID-19 pneumonia (5.06%; 95% CI: 4.5-5.6), required hospitalization (84.08%; 95% CI: 80.0-87.4), a proportion that nearly quadrupled that of COVID-19 pregnant women admitted without pneumonia (24.26%; 95% CI: 22.8-25.7%). The case-fatality rate of COVID-19 pregnant women with pneumonia was ten-fold of that of pregnant women without pneumonia (11.67% [95% CI:8.8-15.3] versus 1.69% [95% CI:1.3-2.2], p<0.0001). Half of pregnant women lived at altitudes above 1185 meters (50.03%, 3724/7444) and 25.02% lived in altitudes above 2187 meters (1863/7774); while 52.4% were born in altitudes above 1567 meters (3903/7444) and 25% in altitudes above 2240 meters (1863/7444). When COVID-19-related symptoms appeared, pregnant women showed a tendency to consult a healthcare center earlier than non-pregnant women (p<0.0001). Although the median time until death of COVID-19 pregnant women was two days longer than non-pregnant patients, no significant differences were found.

Controlling for age and altitude of residence and origin, pregnancy was independently associated with a 15% higher probability of COVID-19 (aPR: 1.15; 47.7% versus 41.4%), a 116% higher probability of its following admission (aPR: 2.169; 22.5% versus 10.4%) and a 127% higher probability of ICU admission (aPR: 2.275; 14.5% versus 6.4%) compared to non-pregnant women without any risk factors. Also, pregnancy was independently associated with 84.2% higher probability of developing COVID-19 pneumonia (aPR: 1.842; 6.4% versus 3.5%) and a 163% higher probability of its following admission (aPR 2.639; 25.3% versus 9.6%). COVID-19 pregnant women seemed to have a higher probability of intubation and a higher case-fatality rate, and those who had pneumonia seemed to have a higher probability of admission and intubation and a lower case-fatality rate compared to non-pregnant women, however no significant differences were found (p=0.279, 0.48, 0.1772, 0.0758 and 0.4182 respectively) (**Table 2**).

**Table 2.** - Adjusted prevalence ratios (aPR) along 95% confidence interval values (CI). Adjusted prevalences (aP) by age and altitude for all patients shown along 95% CI’s. HIghlighted: statistically significant results at p<0.001. n = number of observations used in the prevalence adjustment analysis.

| Outcomes | Pregnant/Non-pregnant women |  | Pregnant women |  | Non-pregnant women |  | n |
| --- | --- | --- | --- | --- | --- | --- | --- |
|  | aPR | 95% CI | aP | 95% CI | aP | 95% CI |  |
| All COVID-19 cases | 1.150 | 1.11-1.19 | 47.7% | 45.9-49.4 | 41.4% | 41.3-41.6 | 334,030 |
| Admission | 2.169 | 2.0-2.34 | 22.5% | 20.7-24.2 | 10.4% | 10.2-10.5 | 138,835 |
| Intubation | 1.360 | 0.71-2.01 | 8.2% | 4.3-12.1 | 6.0% | 5.6-6.4 | 14,921 |
| ICU admission | 2.275 | 1.54-3.0 | 14.5% | 9.9-19.0 | 6.4% | 5.9-6.8 | 14,921 |
| Case-fatality rate | 1.178 | 0.68-1.67 | 2.5% | 1.4-3.5 | 2.1% | 2.0-2.2 | 138,835 |
| Subset: COVID-19 pneumonia | 1.842 | 1.56-2.13 | 6.4% | 5.4-7.4 | 3.5% | 3.4-3.5 | 334,023 |
| Admission | 1.055 | 0.98-1.13 | 73.3% | 67.8-78.7 | 69.5% | 68.7-70.3 | 11,778 |
| Intubation | 1.730 | 0.92-2.54 | 16.5% | 8.9-24.1 | 9.5% | 8.8-10.2 | 8,281 |
| ICU admission | 2.639 | 1.76-3.51 | 25.3% | 17.1-33.5 | 9.6% | 8.9-10.2 | 8,281 |
| Case-fatality rate | 0.853 | 0.5-1.21 | 15.8% | 9.2-22.4 | 18.6% | 17.9-19.3 | 11,778 |

In **Figure 1–A**, we show that the probability of COVID-19 increased with age for both pregnant and non-pregnant women. However, it was always higher in pregnant women, with those over 35 years found with a 50% or higher probability. The chance of developing pneumonia also increased with age for both groups and never exceeded the 13.7% in pregnant women and 7.5% in non-pregnant women (**Figure 1–B**). Hospitalization curves had inverse trends, diminishing for pregnant and increasing for non-pregnant patients. However, the 95% confidence bands for COVID-19 pneumonia overlapped in women older than 45 years (**Figure 1 – C**). The mean chance of intubation for pregnant women over 30 years old was higher compared to non-pregnant women in SARS-CoV-2 infection and over 25 years old in COVID-19 pneumonia, however no significant differences were found for all ages (p>0.05) (**Figure 2 – A-B**). The probability of ICU admission in all COVID-19 cases was higher in pregnant women older than 29 years old and women older than 25 years old in COVID-19 pneumonia compared to non-pregnant women (**Figure 2 – C-D**). Consistent with the adjusted prevalence ratio found, no significant differences were found for case-fatality rates (**Figure 2 – E-F**).

**Image 1.**
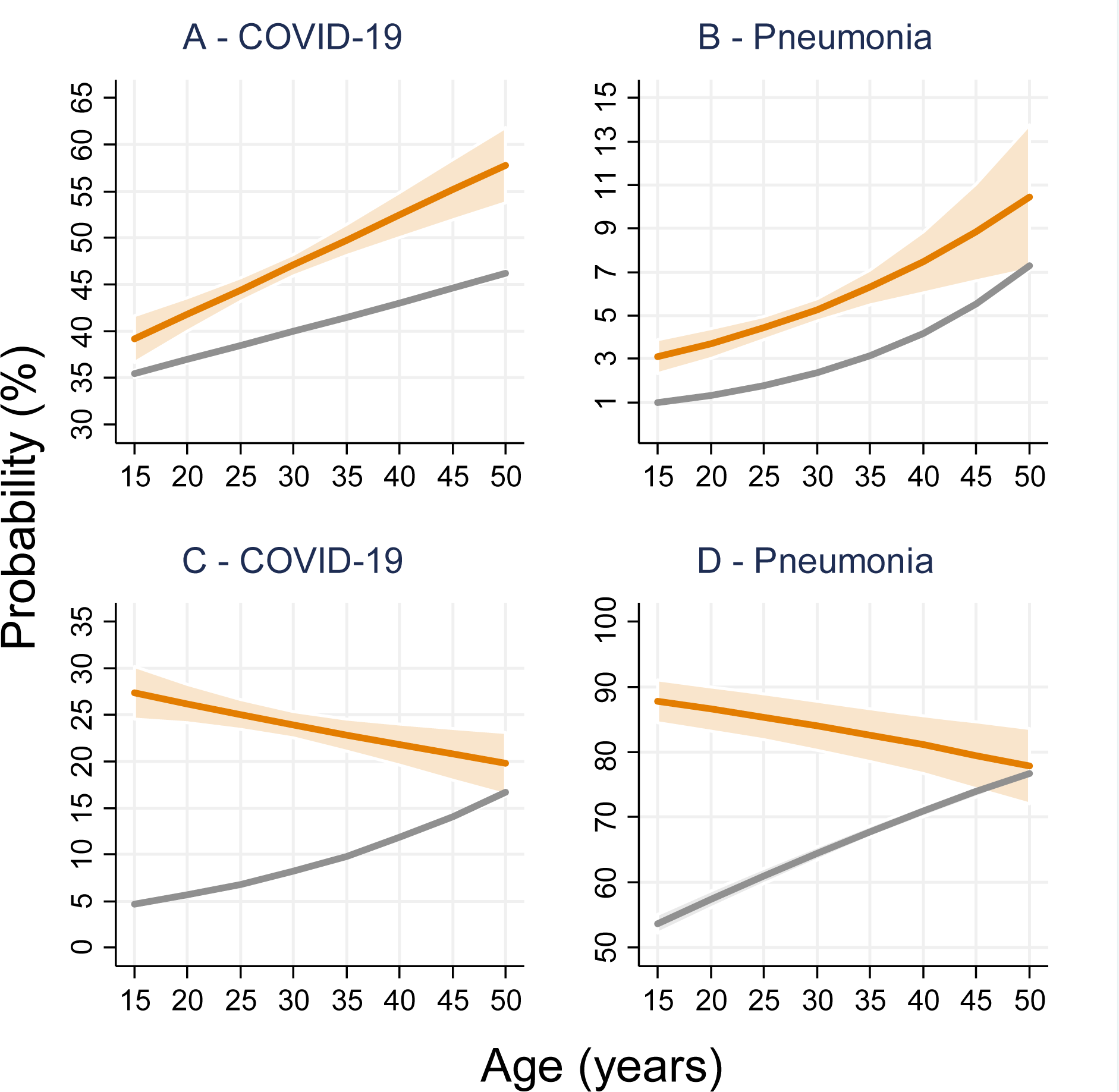
– Mild and moderate COVID-19 and the subset of COVID-19 pneumonia along ages. Gray: Nonpregnant women. Orange: Pregnant women. Non-overlapping confidence bands represent statistically significant differences at the p<0.05 level.

**Image 2.**
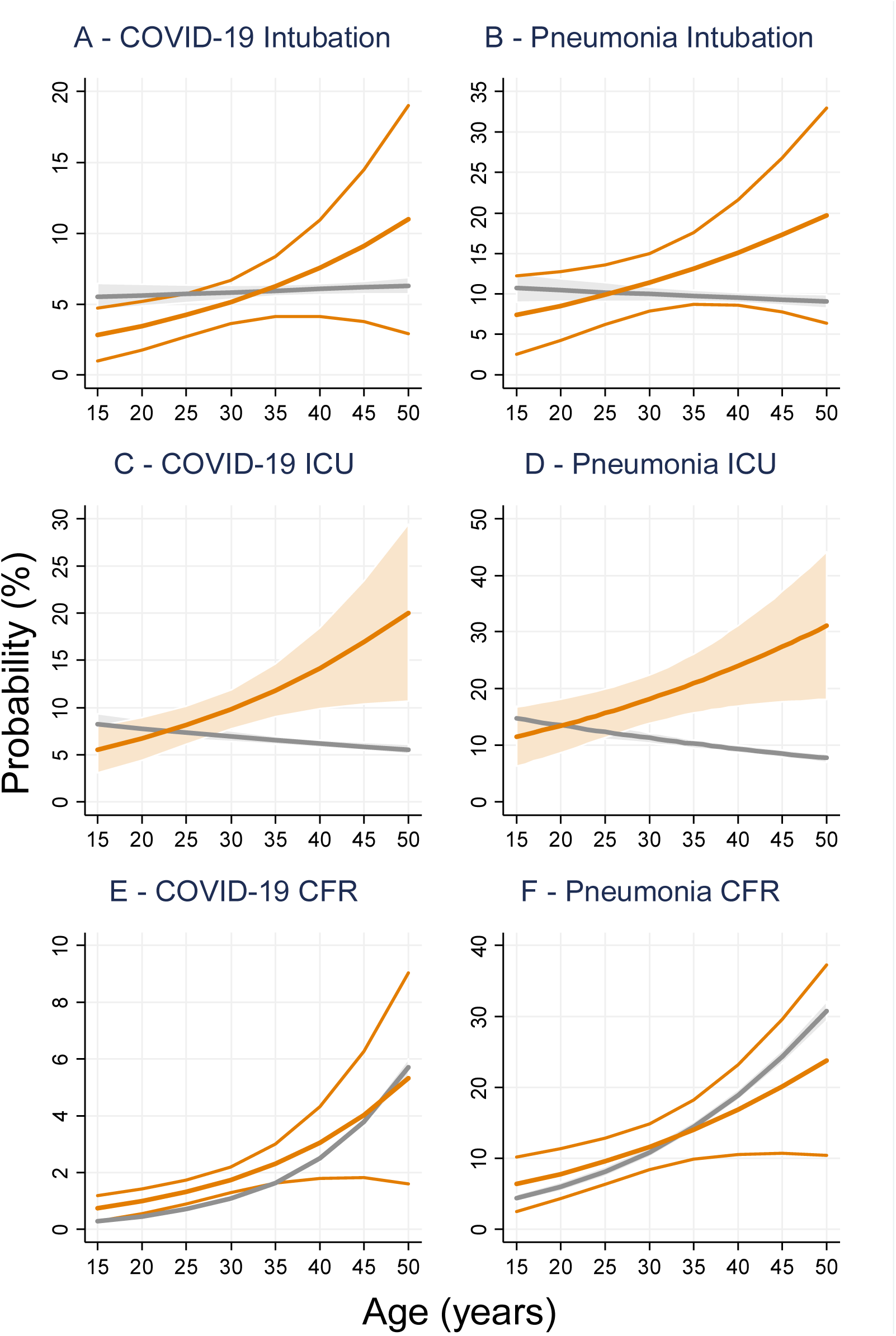
– Severe COVID-19 and the subset of COVID-19 pneumonia along ages. Gray: Non-pregnant women. Orange: Pregnant women. Non-overlapping confidence bands represent statistically significant differences at the p<0.05 level.

As for altitude, the probability trend of COVID-19 (all cases) registered a decreasing trend with pregnant women who resided below 1,750 meters at increased risk and COVID-19 pneumonia for those who resided below 2750 meters (**Figure 3 – A-B**). The chance of admission also decreased with increasing altitudes and was consistently higher in pregnant women (**Figure 3 – C-D**). Pregnant women had the same chance of intubation than no-pregnant across all altitudes (p>0.05), (**Figure 4 – A-B**). The mean probability of ICU admission decreased with altitude but was significantly higher in pregnant women with COVID-19 who lived above 750 meters and all altitudes above sea level in pneumonia cases, compared to non-pregnant women (**Figure 4 – C-D**). In contrast with the adjusted prevalence ratio, the case-fatality rate was found to be lower in pregnant women with COVID-19 who lived above 1000 meters and lower in all pregnant women with pneumonia (**Figure 4 – E-F**).

**Image 3.**
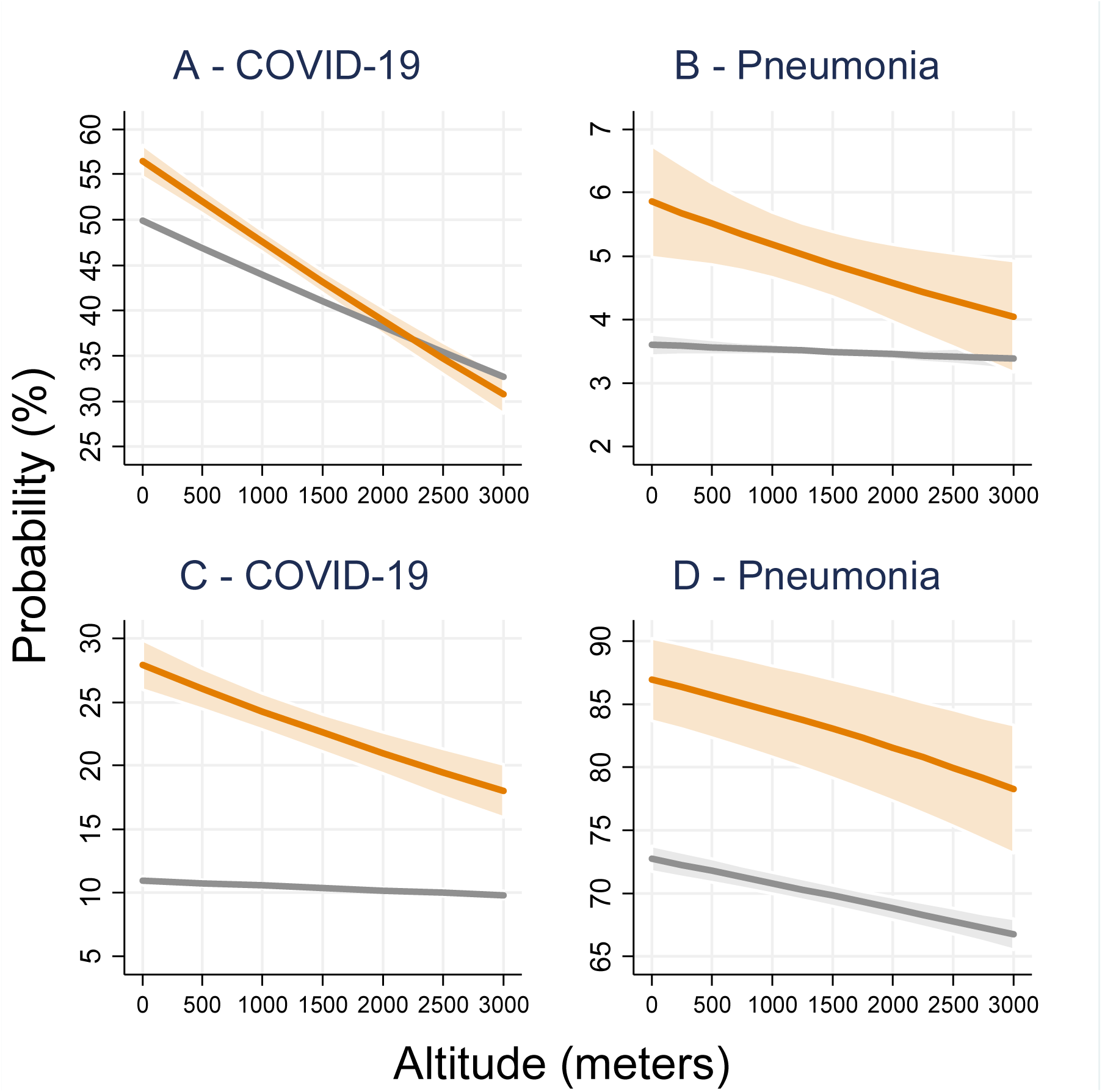
– Mild and moderate COVID-19 and the subset of COVID-19 pneumonia along altitudes. Gray: Nonpregnant women. Orange: Pregnant women. Non-overlapping confidence bands represent statistically significant differences at the p<0.05 level.

**Image 4.**
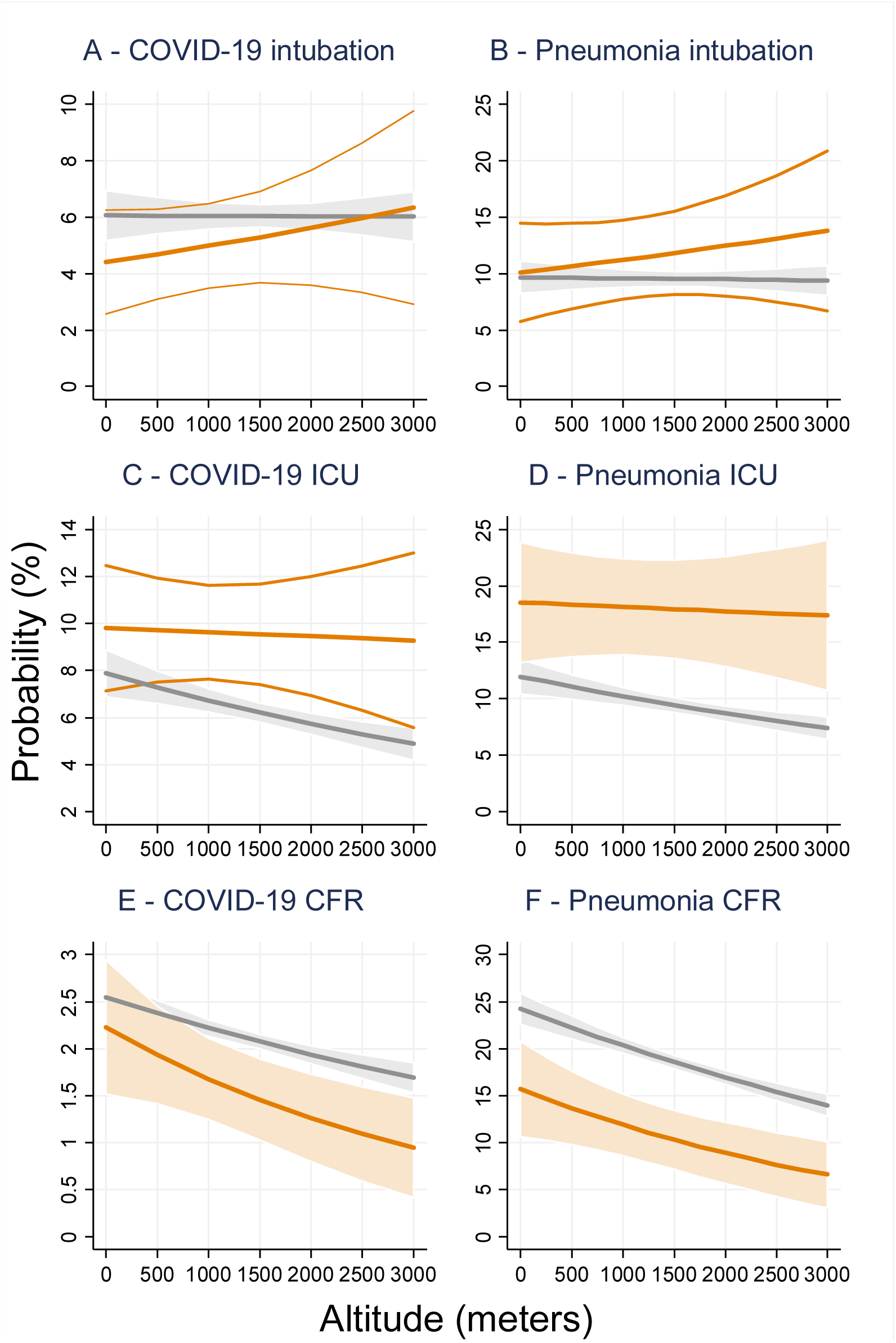
– Severe COVID-19 and the subset of COVID-19 pneumonia along altitudes. Gray: Non-pregnant women. Orange: Pregnant women. Non-overlapping confidence bands represent statistically significant differences at the p<0.05 level.

## Discussion

In the Mexican population, pregnancy was associated with a higher probability of COVID-19 and its following hospitalization and ICU admission; and pneumonia and its following ICU admission compared to non-pregnant women of reproductive age.

Pregnant women displayed a seemingly lower mean probability of intubation until 25-30 years old and a significant lower risk of ICU admission until 25-30 years old than non-pregnant women. With increasing altitudes, pregnant women displayed both a lower probability of COVID-19 and its corresponding case-fatality rates. The data so far does not allow for compelling conclusions about intubation differences according to pregnancy status (**Figure 1-4**).

Pregnant women with influenza, SARS-CoV or MERS-CoV have shown more complications and lethality. During H1N1 Influenza, hospitalization and ICU admission rates were higher compared to the general population. Hospital admission was especially higher in pregnant women in the second half of pregnancy and 2-64% were severe cases (defined as ICU admission or maternal death). Among pregnant women admitted to the ICU, the maternal mortality rate was 0-33% **(13)**. Pregnant women with SARS-CoV infection showed a higher probability of intubation and ICU admission compared to nonpregnant women, in addition to an increased risk of kidney failure and disseminated intravascular coagulopathy. The case fatality rate in pregnant patients with this infection was reported at 25% **(14)**. MERS-CoV infection during pregnancy was only reported in few patients (n=12). Seven of them required admission to the ICU, 3 died, of which 2 contracted MERS-CoV during the third trimester and 1 during the second trimester **(15-16)**.

A systematic review of 538 pregnant women with COVID-19 identified that, according to Wu and McGoogan’s **(17)** classification of disease severity, 86.1% had mild disease, 15.3% severe disease and 1.4% critical disease; while only 3.0% of cases were reported as ICU admissions **(18)**. Although the Mexican dataset did not allow for a standard definition of COVID-19 pneumonia, some parallels could be drawn. Mild cases, defined as those who were treated as outpatients, accounted for 75.74% of all pregnancies with COVID-19 and 15.92% of all pregnancies with pneumonia without other risk factors recorded. While “severe” cases (in which the clinician identified non-reassuring signs and symptoms related or not to pregnancy) that were more likely to be admitted represented 24.26% of pregnant women with COVID-19 and 84.08% in the subset of pregnant women with COVID-19 pneumonia. The rate of pregnant women with COVID-19 “critical” cases (Those who required ICU admission) stood at 9.63%; however, pregnant patients with COVID-19 pneumonia had a higher rate of ICU admission (18.10%) followed by an overall COVID-19 1.69% case-fatality rate that increased by several orders of magnitude if pneumonia was also present (11.67%).

The largest case series of women of reproductive age (15-45 years) reported by the Centers of Disease Control and Prevention **(19)** found that among the 9% of pregnant women (8207/91412), 31.5% required hospitalization, 1.5% needed ICU support and 0.2% died. This represents up to 46.4% additional hospitalizations in comparison to the Mexican series. The difference in the percentages could be due to the lower proportion (2.23%) of pregnancies in women of reproductive age (15-49 years) in our study, in addition to characteristics inherent to their population or their health system.

The Angiotensin-Converting Enzyme 2 (ACE2) is a receptor on the cell surface to which binds SARS-CoV-2 for entering inside it. During pregnancy, ACE2 activity can increase up to twofold, being the placenta and the uterus the most important sources of ACE2 **(20)**. This increased activity responds to the need of blood pressure regulation in pregnancy **(21)**. Also, ACE2 expression is critical for both an adequate pregnancy progression and fetal development, as its deficiency relates to both impaired gestational weight and fetal growth restriction **(22)**. Lower estrogen activity, a down regulator of ACE2 expression, may contribute in its increased activity seen in pregnancy. 17β-estradiol (E2) is physiologically found in women during pregnancy and has been shown to reduce ACE2 mRNA in differentiated normal human bronchial epithelial cells (NHBE), which does not necessarily mean a reduction of surface ACE2 protein **(23)**. However, these reports could be important to understand why pregnant women did not have a higher risk of mortality from SARS-CoV-2. Therefore, the increased ACE2 pool could render pregnant women more susceptible to developing COVID-19 pneumonia. However, it remains unclear how the complexity of the immune system that renders pregnant women more susceptible to viral, bacterial, parasitic and autoimmune disorders interact with the inflammatory response associated with severe COVID-19 cases **(24)**.

Regarding altitude, we found that in pregnant women altitude influenced the trend of COVID-19, development of pneumonia, admission, intubation, and maternal case-fatality rates (Figure 3-4). The main significant relationship was found for pregnant women with COVID-19 compared to their baseline probability (of non-pregnant women) as all saw falling rates that were higher that of non-pregnant women only below 1750 meters. The admission trend was always higher for pregnant women but tended to decay with higher altitudes (**Figure 3**). Finally, the last difference was seen at case-fatality rates, finding that pregnant women are at lower CFR above 750 meters (**Figure 4 – E**). This paper confirms that predictive models could be used to monitor populations at risk through probability plots **(25)**. This approach has the advantage of performing a regression on the continuous spectrum of altitudes instead of assigning a static value for altitude ranges, which could introduce cluster effect and difficult the interpretation of results.

Arias-Reyes first observed less COVID-19 rates in high-altitude dwellers compared to populations living below 3,000 meters. The authors related this association with a possible lower ACE2 expression in inhabitants of high altitude **(26)**. This observation was further observed by the same authors in patients living below 1000 meters **(27)**. In Peru, researchers also found that COVID-19 rates and its subsequent mortality fall logarithmically with increasing altitudes **(10)**. Of note, hypobaric hypoxia is classically described to start at 1500 meters due to a reduced alveolar diffusion pressure and reduced mass transport **(28)**. As the hypobaric hypoxia found at high altitudes upregulates the hypoxia inducible factor 1 alpha (HIF1-a), ACE2 downregulation ensues **(29)**. Under experimental conditions, TMPRSS2 is also downregulated via HIF-signaling **(30)**. As these membrane enzymes represent the gate and the doorkeeper of viral cell entry **(31)**, patients already residing at high altitudes would benefit with a reduced chance of SARS-CoV-2 infection. Although both SARS-CoV and SARS-CoV-2 share the same entry receptor (ACE2), historical observations under hypoxia in SARS-CoV were not feasible due to the geographical distribution of the disease that was not reported in high-altitude areas within the affected countries (**Image 5**).

**Image 5.**
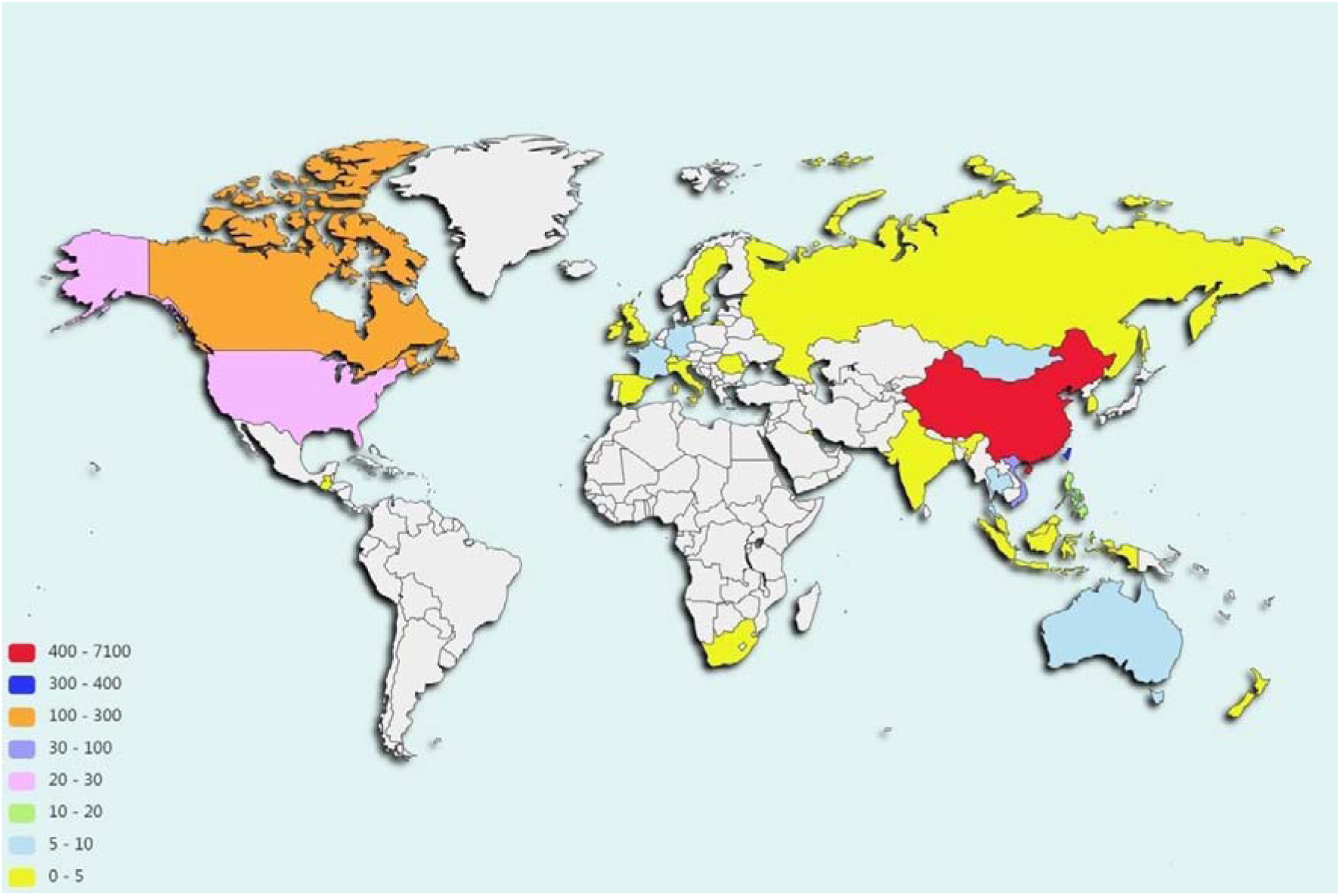
– Geographical distribution of SARS-CoV.

Apart from ACE2, erythropoietin (EPO) was also suggested as a putative factor protecting human population at high altitude from COVID-19 **(32)**. Hypobaric hypoxia induces 2−4-fold increases in serum levels of EPO in permanent citizens at high altitude compared to lowlanders **(33)**. However, independently from altitude, EPO levels rise during pregnancy, especially during the first trimester, to 2-4 times the concentration compared with nonpregnant control subjects **(34)**. EPO is certainly the essential hormone for the stimulation of red blood cells production and reticulocytosis in the bone marrow (35-36). This aspect is central in the infectious scenario of COVID-19, as EPO may help restoring hemoglobin levels and improve oxygen delivery to the tissues. In addition, EPO also promotes adaptive cellular responses to hypoxic challenges and tissue-damaging insults in non-hematopoietic tissues **(37)**. Non-erythroid effects of EPO include the activation of anti-apoptotic, anti-oxidative, anti-cytotoxic, and anti-inflammatory mechanisms **(38)** in several tissues, including the brain, lung, kidney, vascular bed, and heart (39-43). As such, EPO may play a key role against generalized tissue damage, multiple organ dysfunction and inflammation induced by SARS-CoV-2 in lowland pregnant women in general, but especially in pregnant women at high-altitude (44-45). In fact, increased levels of EPO with altitude could explain, partially, the altitude-related gradual decreasing tendency of pregnant women to acquire SARS-CoV-2, develop COVID-19 pneumonia, hospitalization, and maternal death observed in our work. For other respiratory viral infections, as influenza, high altitude rates are only 30% lesser, **(46)** but for COVID-19 in Peru they are 350% lesser, being the decrease in ECA-2 expression related to hypobaric hypoxia probably the reason **(10)**.

Limitations of the study included that the dataset did not record the trimester of pregnancy, therefore we couldn’t determined the clinical reason for admission to regular hospitalization or ICU or whether the patient was admitted preventively and subsequently worsened their respiratory parameters requiring intubation. Because of this, it remains unclear how SARS-CoV-2 infection in the first trimester affects women and fetal stability across the course of pregnancy, despite that SARS-CoV-2 transplacental transmission has been reported previously **(47)**. Also, pregnant women’s physiology varies across gestational age and might interact with the hypobaric hypoxia seen at high altitudes. In order to isolate the effects of pregnancy, we only included women with no other recorded risk factors in the analysis, so the burden of chronic diseases such as hypertension and obesity remains to be scrutinized.

## Conclusion

Pregnancy was associated with a higher probability of SARS-CoV-2 infection, COVID-19 pneumonia and need for hospitalization, but not for case fatality rates. Furthermore, the still developing information on this new coronavirus limits the understanding of its possible effect during pregnancy. However, our results suggest that as the altitude increase, the risk of SARS-CoV-2 infection decreases. Despite, various aspects of this virus are yet unknown, ACE2 and EPO may play important role in this phenomenon. Moreover, our data suggests that knowledge of the mechanisms of physiological adjustment to hypoxia may help to better understand COVID-19. At the same time, this knowledge can lead to the development of treatment strategies specifically focused on the population of pregnant women.

## Data Availability

Processed data files used in the present study are available at request. The authors follow the STrengthening the Reporting of OBservational studies in Epidemiology (STROBE) statement for data collection, analysis and reporting.

https://www.gob.mx/salud/documentos/datos-abiertos-152127

## Declaration of conflicts of interest

The authors declare that no financial, academic or institutional conflicts of interest that affected the conception, elaboration, execution and processing of this paper.

## Funding

The paper has not received any kind of funding aid from public agencies, the commercial sector or non-profit entities.

## Notes

### Competing Interest Statement

The authors have declared no competing interest.

### Author Declarations

This study was waived from revision by the Universidad Peruana Cayetano Heredia IRB with registry number SIDISI 202658.

## References

1. Goodnight WH, Soper DE. Pneumonia in pregnancy. Critical Care Medicine. 2005 Oct;33(Supplement):S390-S397.

2. Mighty He. Acute Respiratory Failure in Pregnancy. Clinical Obstetrics and Gynecology. 2010 Jun;53(2):360–8.

3. Jamieson D, Theiler R, Rasmussen S. Emerging Infections and Pregnancy. Emerg Infect Dis. 2006;12(11):1638–43

4. World Health Organization. Guidelines for the global surveillance of severe acute respiratory 361 syndrome (SARS). Geneva: World Health Organization; October 2004. Accessed on June12, 2020. 363 4.

5. World Health Organization, Regional Office for Eastern Mediterranean. MERS situation 364 update. Geneva: World Health Organization; January 2020. Accessed on June 12, 2020.

6. Centers for Disease Control and Prevention. 2009 H1N1 Pandemic (H1N1pdm09 virus) [Internet]. Centers for Disease Control and Prevention; 2019 [cited 2020 Jun11]. Available from:https://www.cdc.gov/flu/pandemic-resources/2009-h1n1-pandemic.html

7. Dawood FS, Iuliano AD, Reed C, Meltzer MI, Shay DK, Cheng P, et al. Estimated global mortality associated with the first 12 months of 2009 pandemic influenza A H1N1 virus circulation: a modelling study. The Lancet Infectious Diseases. 2012 Sep;12(9):687–95.

8. WHO Coronavirus Disease (COVID-19) Dashboard [Internet]. Covid19.who.int. 2020 [cited 16 August 2020]. Available from: https://covid19.who.int/

9. Centers of disease control and prevention. If You Are Pregnant, Breastfeeding, or Caring for Young Children. 2020. Available from: https://www.cdc.gov/coronavirus/2019-ncov/need-extra-precautions/pregnancy-breastfeeding.html

10. Accinelli RA, Leon-Abarca JA. At high altitude COVID-19 is less frequent: The experience of Peru. [Article in Spanish]. Arch Bron. 2020 Jul 16.

11. Secretaría de Salud. DatosAbiertos - Dirección General de Epidemiología. Gobierno de México. [2020 Aug 9; 2020 Aug 10]. Available from: https://www.gob.mx/salud/documentos/datos-abiertos-152127

12. Von Elm E, Altman DG, Egger M, Pocock SJ, Gøtzsche PC, Vandenbroucke JP. The Strengthening the Reporting of Observational Studies in Epidemiology (STROBE) statement: guidelines for reporting observational studies. Annals of internal medicine. 2007 Oct 16;147(8):573–7.

13. Meijer WJ, van Noortwijk AG, Bruinse HW, Wensing AM. Influenza virus infection in pregnancy: a review. ActaObstetGynecol Scand. 2015 Aug;94(8):797–819.

14. Wong SF, Chow KM, Leung TN, Ng WF, Ng TK, Shek CC, et al. Pregnancy and perinatal outcomes of women with severe acute respiratory syndrome. American Journal of Obstetrics and Gynecology. 2004 Jul;191(1):292–7.

15. Alfaraj SH, Al-Tawfiq JA, Memish ZA. Middle East Respiratory Syndrome Coronavirus (MERS-CoV) infection during pregnancy: Report of two cases & review of the literature. Journal of Microbiology, Immunology and Infection. 2019 Jun;52(3):501–3.

16. Jeong SY, Sung SI, Sung J, Ahn SY, Kang E, Chang YS, et al. MERS-CoV Infection in a Pregnant Woman in Korea. J Korean Med Sci. 2017;32(10):1717.

17. Wu Z, McGoogan JM. Characteristics of and important lessons from the coronavirus disease 2019 (COVID-19) outbreak in China: summary of a report of 72 314 cases from the Chinese Center for Disease Control and Prevention. Jama. 2020 Apr 7;323(13):1239–42.

18. Huntley BJ, Huntley ES, Di Mascio D, Chen T, Berghella V, Chauhan SP. Rates of Maternal and Perinatal Mortality and Vertical Transmission in Pregnancies Complicated by Severe Acute Respiratory Syndrome Coronavirus 2 (SARS-Co-V-2) Infection: A Systematic Review. Obstetrics & Gynecology. 2020 Jun 9.

19. Ellington S, Strid P, Tong VT, Woodworth K, Galang RR, Zambrano LD, Nahabedian J, Anderson K, Gilboa SM. Characteristics of Women of Reproductive Age with Laboratory-Confirmed SARS-CoV-2 Infection by Pregnancy Status — United States, January 22–June 7, 2020. MMWR Morb Mortal Wkly Rep 2020;69:769–775.

20. Levy A, Yagil Y, Bursztyn M, Barkalifa R, Scharf S, Yagil C. ACE2 expression and activity are enhanced during pregnancy. American Journal of Physiology-Regulatory, Integrative and Comparative Physiology. 2008 Dec;295(6):R1953-61.

21. Zhao X, Jiang Y, Zhao Y, Xi H, Liu C, Qu F, Feng X. Analysis of the susceptibility to COVID-19 in pregnancy and recommendations on potential drug screening. European Journal of Clinical Microbiology & Infectious Diseases. 2020 Apr 23:1.

22. Bharadwaj MS, Strawn WB, Groban L, Yamaleyeva LM, Chappell MC, Horta C, Atkins K, Firmes L, Gurley SB, Brosnihan KB. Angiotensin-converting enzyme 2 deficiency is associated with impaired gestational weight gain and fetal growth restriction. Hypertension. 2011 Nov;58(5):852–8.

23. Stelzig KE, Canepa-Escaro F, Schiliro M, Berdnikovs S, Prakash YS, Chiarella SE. Estrogen regulates the expression of SARS-CoV-2 receptor ACE2 in differentiated airway epithelial cells. American Journal of Physiology-Lung Cellular and Molecular Physiology. 2020.

24. Berghella V. Coronavirus disease 2019 (COVID-19): Pregnancy issues. UpToDate. Internet. 2020.

25. Leon-Abarca JA. Modeling the progression of SARS-CoV-2 infection in patients with COVID-19 risk factors through predictive analysis. medRxiv. 2020 Jul 19.

26. Arias-Reyes C, Zubieta-DeUrioste N, Poma-Machicao L, Aliaga-Raudan F, Carvajal-Rodriguez F, Dutschmann M, Schneider-Gasser E, Zubieta-Calleja G,Soliz J. Does the pathogenesis of SAR-CoV-2 virus decrease at high-altitude?. Respiratory Physiology & Neurobiology. 2020 Apr 22:103443.

27. Arias-Reyes C, Carvajal-Rodriguez F, Poma-Machicao L, Aliaga-Raduan F, Marques DA, DeUrioste NZ, Accinelli R, Schneider-Gasser EM, Zubieta-Calleja G, Dutschmann M, Soliz J. Decreased incidence, virus transmission capacity, and severity of COVID-19 at altitude on the American continent. medRxiv. 2020 Aug 1.

28. Böning D. Altitude and hypoxia training-a short review. International journal of sports medicine. 1997 Nov;18(08):565–70.

29. Zhang R, Wu Y, Zhao M, Liu C, Zhou L, Shen S, Liao S, Yang K, Li Q, Wan H. Role of HIF-1α in the regulation ACE and ACE2 expression in hypoxic human pulmonary artery smooth muscle cells. American Journal of Physiology-Lung Cellular and Molecular Physiology. 2009 Oct;297(4):L631-40.

30. Fernandez EV, Reece KM, Ley AM, Troutman SM, Sissung TM, Price DK, Chau CH, Figg WD. Dual targeting of the androgen receptor and hypoxia-inducible factor 1α pathways synergistically inhibits castration-resistant prostate cancer cells. Molecular pharmacology. 2015 Jun 1;87(6):1006–12.

31. Hoffmann M, Kleine-Weber H, Schroeder S, Krüger N, Herrler T, Erichsen S, Schiergens TS, Herrler G, Wu NH, Nitsche A, Müller MA. SARS-CoV-2 cell entry depends on ACE2 and TMPRSS2 and is blocked by a clinically proven protease inhibitor. Cell. 2020 Mar 5.

32. Soliz J, Schneider-Gasser EM, Arias-Reyes C, Aliaga-Raduan F, Poma-Machicao L, Zubieta-Calleja G, Furuya WI, Trevizan-Baú P, Dhingra RR, Dutschmann M. Coping with hypoxemia: Could erythropoietin (EPO) be an adjuvant treatment of COVID-19?. Respiratory Physiology & Neurobiology. 2020 Jun 6:103476.

33. Beall CM. Detecting natural selection in high-altitude human populations. Respiratory physiology & neurobiology. 2007 Sep 30;158(2-3):161-71.

34. Sienas L, Wong T, Collins R, Smith J. Contemporary uses of erythropoietin in pregnancy: a literature review. Obstetrical & gynecological survey. 2013 Aug 1;68(8):594–602.

35. Martin D, Windsor J. From mountain to bedside: understanding the clinical relevance of human acclimatisation to high-altitude hypoxia. Postgraduate medical journal. 2008 Dec 1;84(998):622–7.

36. Storz JF, Moriyama H. Mechanisms of hemoglobin adaptation to high altitude hypoxia. High altitude medicine & biology. 2008 Jun 1;9(2):148–57.

37. Juul S, Felderhoff-Mueser U. Epo and other hematopoietic factors. InSeminars in Fetal and Neonatal Medicine 2007 Aug 1 (Vol. 12, No. 4, pp. 250-258). WB Saunders.

38. Peng B, Kong G, Yang C, Ming Y. Erythropoietin and its derivatives: from tissue protection to immune regulation. Cell Death & Disease. 2020 Feb 3;11(2):1–2.

39. Arcasoy MO. Non-erythroid effects of erythropoietin. Haematologica. 2010;95: 1803-1805.

40. Kakavas S, Demestiha T, Vasileiou P, Xanthos T. Erythropoetin as a novel agent with pleiotropic effects against acute lung injury. European journal of clinical pharmacology. 2011 Jan 1;67(1):1–9.

41. Zhang Y, Zhu X, Huang X, Wei X, Zhao D, Jiang L, Zhao X, Du Y. Advances in Understanding the Effects of Erythropoietin on Renal Fibrosis. Frontiers in Medicine. 2020 Feb 21;7:47.

42. Bitto A, Irrera N, Pizzino G, Pallio G, Mannino F, Vaccaro M, Arcoraci V, Aliquò F, Minutoli L, Colonna MR, Galeano MR. Activation of the EPOR-β common receptor complex by cibinetide ameliorates impaired wound healing in mice with genetic diabetes. Biochimica et Biophysica Acta (BBA)-Molecular Basis of Disease. 2018 Feb 1;1864(2):632–9.

43. Xu B, Dong GH, Liu H, Wang YQ, Wu HW, Jing H. Recombinant human erythropoietin pretreatment attenuates myocardial infarct size: a possible mechanism involves heat shock Protein 70 and attenuation of nuclear factor-kappaB. Annals of Clinical & Laboratory Science. 2005 Apr 1;35(2):161–8.

44. Chen N, Zhou M, Dong X, Qu J, Gong F, Han Y, Qiu Y, Wang J, Liu Y, Wei Y, Yu T. Epidemiological and clinical characteristics of 99 cases of 2019 novel coronavirus pneumonia in Wuhan, China: a descriptive study. The Lancet. 2020 Feb 15;395(10223):507–13.

45. Yang X, Yu Y, Xu J, Shu H, Liu H, Wu Y, Zhang L, Yu Z, Fang M, Yu T, Wang Y. Clinical course and outcomes of critically ill patients with SARS-CoV-2 pneumonia in Wuhan, China: a single-centered, retrospective, observational study. The Lancet Respiratory Medicine. 2020 Feb 24.

46. Tinoco YO, Azziz-Baumgartner E, Uyeki TM et al. Burden of influenza in 4 ecologically distinct regions of Peru: household active surveillance of a community cohort, 2009–2015. Clinical Infectious Diseases. 2017 Oct 16;65(9):1532–41.

47. Vivanti A, Vauloup-Fellous C, Prevot S, Zupan V, Suffee C, Do Cao J, Benachi A, De Luca D. Transplacental transmission of SARS-CoV-2 infection. Nat Commun. 2020;11:3572.

